# Quantified Flu: an individual-centered approach to gaining sickness-related insights from wearable data

**DOI:** 10.1101/2021.03.10.21252242

**Authors:** Bastian Greshake Tzovaras, Enric Senabre, Karolina Alexiou, Lukaz Baldy, Basille Morane, Ilona Bussod, Melvin Fribourg, Katarzyna Wac, Gary Wolf, Mad Ball

## Abstract

**Background:** Wearables have been used widely for monitoring health in general, and recent research results show that they can be used for predicting infections based on physiological symptoms. So far, the evidence has been generated in large, population-based settings. In contrast, the Quantified Self and Personal Science communities are composed of people interested in learning about themselves individually using their own data, often gathered via wearable devices.

**Objective:** We explore how a co-creation process involving a heterogeneous community of personal science practitioners can develop a collective self-tracking system to monitor symptoms of infection alongside wearable sensor data.

**Methods:** We engaged in a co-creation and design process with an existing community of personal science practitioners, jointly developing a working prototype of an online tool to perform symptom tracking. In addition to the iterative creation of the prototype (started on March 16, 2020), we performed a netnographic analysis, investigating the process of how this prototype was created in a decentralized and iterative fashion.

**Results:** The Quantified Flu prototype allows users to perform daily symptom reporting and is capable of visualizing those symptom reports on a timeline together with the resting heart rate, body temperature, and respiratory rate as measured by wearable devices. We observe a high level of engagement, with over half of the 92 users that engaged in the symptom tracking becoming regular users, reporting over three months of data each. Furthermore, our netnographic analysis highlights how the current Quantified Flu prototype is a result of an interactive and continuous co-creation process in which new prototype releases spark further discussions of features and vice versa.

**Conclusions:** As shown by the high level of user engagement and iterative development, an open co-creation process can be successfully used to develop a tool that is tailored to individual needs, decreasing dropout rates.

## Introduction

Patient- or participant-led research has been suggested to improve self-management capabilities [1] and provide ways to generate otherwise undone science [2], [3]. A particular subtype of participant-led research is *personal science*, which involves the use of empirical methods by individuals to pursue personal health questions [4]. Personal science is a distinct category of citizen science that has emerged from the Quantified Self community and its efforts to advance participant-led research [5], [6]. In personal science, practitioners almost always take the lead in all stages of the research process by definition [4]. Due to this high level of individual engagement and tailoring to individuals’ interests, personal science has the potential to deliver novel insights relevant for its practitioners [7] that can lead to an improved sense of agency and quality of life [8]. Furthermore, the insights and self-expertise generated by these types of participant-led processes have potential relevance for professional, scientific research, both topically as a source of ideas and methodologically as a source of tools, analytical approaches, and workflows [9].

Wearable devices – from wristbands to smartwatches and other personalized, miniaturized on- and around-body devices – are frequently used by self-trackers. These devices are becoming more and more common and are used for a wide spectrum of well-being, fitness, and health-related purposes [10]. This is further facilitated by the fact that the number of sensors used in those devices is growing rapidly: In addition to accelerometers and gyroscopes to track physical activity, sensors to measure physiological signals such as heart rate, body temperature, respiratory rate, and blood oxygen saturation, that may correspond to health/sickness state of the human body [11], are now frequently found in wearables as well [12], [13]. Consequently, even outside the realms of *personal science* wearables, have long been seen as promising tools to facilitate health-related monitoring and enable personalized medicine [14], [15], and have been proposed or used to monitor conditions as diverse as cardiovascular disease [16], [17], Alzheimer’s [18] and graft-versus-host disease [19].

In response to the COVID-19 pandemic, interest in using wearable technology for infection prediction and surveillance has increased [20]–[22]. Anecdotal reports from self-trackers suggested that wearables may provide evidence of COVID-19 infection [23]. During the first year of the COVID-19 pandemic, a small number of studies appeared, highlighting that wearable devices, often along with self-reported symptoms, might indeed be used for the early detection of COVID-19 infections and to assess physiological symptoms [24]–[27]. The majority of these studies take a crowdsourcing-based approach – in which participants are invited to contribute by providing their own wearable data along with regular symptom reports and COVID-19 test results – as the main way of engaging individuals. The goal of the data collection process in these studies is to create big data sets to interrogate.

In contrast, there have been only limited efforts that try to engage personal scientists in co-creating such symptom tracking efforts. Personal science practices are largely done in isolation, and the Quantified Self movement shows consequently limited knowledge accumulation so far [8]. To fill this gap, we present a case study of *Quantified Flu* (QF), a project co-created by a community of personal science practitioners in response to the COVID-19 pandemic.

The goals of this work are two-fold: First, we document the contrasting co-creation approach of QF with its focus on personal science rather than large-scale research. To this end, we use netnographic methods to document how the co-creation process developed and generated a citizen science platform prototype over a relatively short period of time. Second, we explore the consequences of the projects’ contrasting co-creation approach and focus on personal science, in particular with respect to the ultimate design of the QF tool as well as its usage.

## Methods

The co-creation process of this study is based on an action research approach [28], simultaneously for developing a useful community resource while also generating shared knowledge about the process. In our case, action research was implemented through the practical work to support the participatory design of a digital platform [29], [30] under open source principles [31], followed by netnographic data collection and analysis to understand its development and usefulness as a co-creation process [32]. For this, all authors except E.S.H. were involved as participants in the co-creation process, in collaboration with the rest of the participants, during the iterative prototyping of the QF platform.

### Community co-creation process

QF started out from a discussion on the monthly *Open Humans* community call at the beginning of the COVID-19 pandemic on March 10, 2020. Open Humans (OH) is a platform for empowering individuals around their personal data, to explore and share research processes for the purposes of education, health, and science in general [33]. The community calls involved 83 individuals so far (until September 03, 2020), and the monthly calls are frequented by a mix of citizen science and personal science practitioners; usually, around ten individuals take part in each call. Following an initial brainstorming, the discussions and planning stages were continued through following community calls and a dedicated communication channel of the OH community Slack [34]. Furthermore, over the evolution of the project, other communities like the *Quantified Self* [35] and the *Open Covid19 Initiative* [36] were engaged and involved in different aspects of the development of the project.

In parallel to 10 additional community calls between March 10 and September 03, 2020, the main coordination tool for the QF project was a specific Slack channel with a total of 146 subscribers and 50 active users over time (33% participation) with different levels of involvement and activity. During this timeframe, this openly accessible channel gathered a total of 844 messages from these users, with a total count of 26,691 words (and 3,917 unique words).

While the planning, coordination, and social aspects of the co-creation process mainly took place on the mentioned project’s Slack channel, the technical collaboration and software development happened through GitHub and the git repository of the QF. Due to the iterative nature of the open source collaboration, no upfront requirement analysis was performed. Instead, the prototyping developed over time according to community discussions by iteratively adding and testing implementations. On GitHub, 7 contributors created a total of 316 commits since March 12 2020, leading to the technical prototype that is outlined below. The source code for the project is available under an open license on GitHub [37].

### Netnographic content analysis

To investigate and analyze the co-creation stages that led to the QF prototype we performed a netnographic analysis of its iterative communication process, similar to previous studies about co-creation in health-related community settings [38]. Netnography is an interpretive research method derived from ethnography, usually applied to social interaction processes in digital channels and platforms, and focused on digital traces of public conversations as analysable data. As a qualitative technique broadly applied to the study of online communities [32], Netnography allows to capture and reflect interactions as an observational, inductive, and unobtrusive approach, while at the same time combining it with participatory methods [39]. In particular, we examine how individuals engaged in the QF Slack channel for the collaborative development of the QF platform as a case study setting [40].

For this part of data collection, one of the researchers (E.S.H.) developed a codebook combining key concepts of co-creation and collaboration in communities of practice (table 1). The codebook was cross-checked for validity by two other authors (B.G.T. and M.B.). Following this, it was applied to the QF Slack channel *post-hoc*, without this specific researcher (E.S.H.) having participated in the previous community discussions.

**Table 1:** Codebook for QF Slack communication messages content analysis

|  |  |  |
| --- | --- | --- |
| **Communities of** | **Socialization** | **Support / coordination:** Parallel messages regarding overall coordination, also personal and empathic support interventions |
| practic<br>e<br>related<br>messag<br>es |  | **Possible collaborations:** Ideas regarding potential collaborators, connection to other organizations or experts who can support or contribute to the project |
|  |  | **Outreach:** Messages related to the visibility of the project, possible dissemination, or alliances for spreading the process |
|  |  | **Off topic:** Non-related messages to any of the previous (for example, about personal issues, or intention to buy wearables, etc) |
| Co-<br>creatio<br>n<br>related<br>messag<br>es | Ideas | **Inspiring / similar initiatives:** Mentions to other COVID-related projects, being developed or known externally |
|  |  | **Covid related:** Links to news or updates regarding the covid pandemic and its evolution |
|  |  | **Mention to tool / wearable:** References to specific wearable for its potential connection to the QF project |
|  |  | **Scientific knowledge / papers:** Mentions or links to studies or publications and elaborated scientific knowledge |
|  | QF<br>Concept | **Goal setting / discussion:** Concept-related interventions about the objectives of the project |
|  |  | **Protocol / tool design:** Mentions to how the protocol and tool should work, or specific aspects of its possible design |
|  |  | **Feature suggestion:** Interventions suggesting specific characteristics or new possible features of the tool |
|  |  | **Pattern / data observation:** Statements regarding the observation of data in relation to the goals or possible functioning of the project |
|  | QF<br>Prototype | **Incremental development / updates:** Messages informing about new implementations, code development of features applied to the prototype |
|  |  | **Technical issues:** Specific technical issues to solve or observations about needed improvements for correct use |
|  |  | **Help testing:** Interventions asking or offering support in testing the tool by community members |
|  |  | **Help developing:** Interventions asking or offering technical support for the development of the tool |

This part of data analysis was used to determine the typology of messages regarding the co-creation of the QF platform, from idea to concept and to prototype [29], as well as other types of messages relevant from a communicational and empathy-needed dialogic process in communities of practice [41]. Each Slack message was assigned up to three top tags, based on the above codebook categories, depending on its text density and characteristics. The researcher’s (E.S.H.) assessments of types and categories of messages were afterward reviewed and discussed by another co-author (B.G.T.), who was actively involved during the analysed co-creation process.

## Results

We present the current prototype of the *Quantified Flu* platform (available at [42]), as a result of the described technical development, before analyzing the co-creation process that led to it.

### Community-based development

A first overview derived from our netnographic analysis of the four main categories of messages interchanged during the co-creation of the QF prototypes on its dedicated Slack channel (March 10 to September 03, 2020) shows a relative balance in the topics of the online messages among the 1,171 message fragments that were annotated (see examples in Table 2).

**Table 2:**
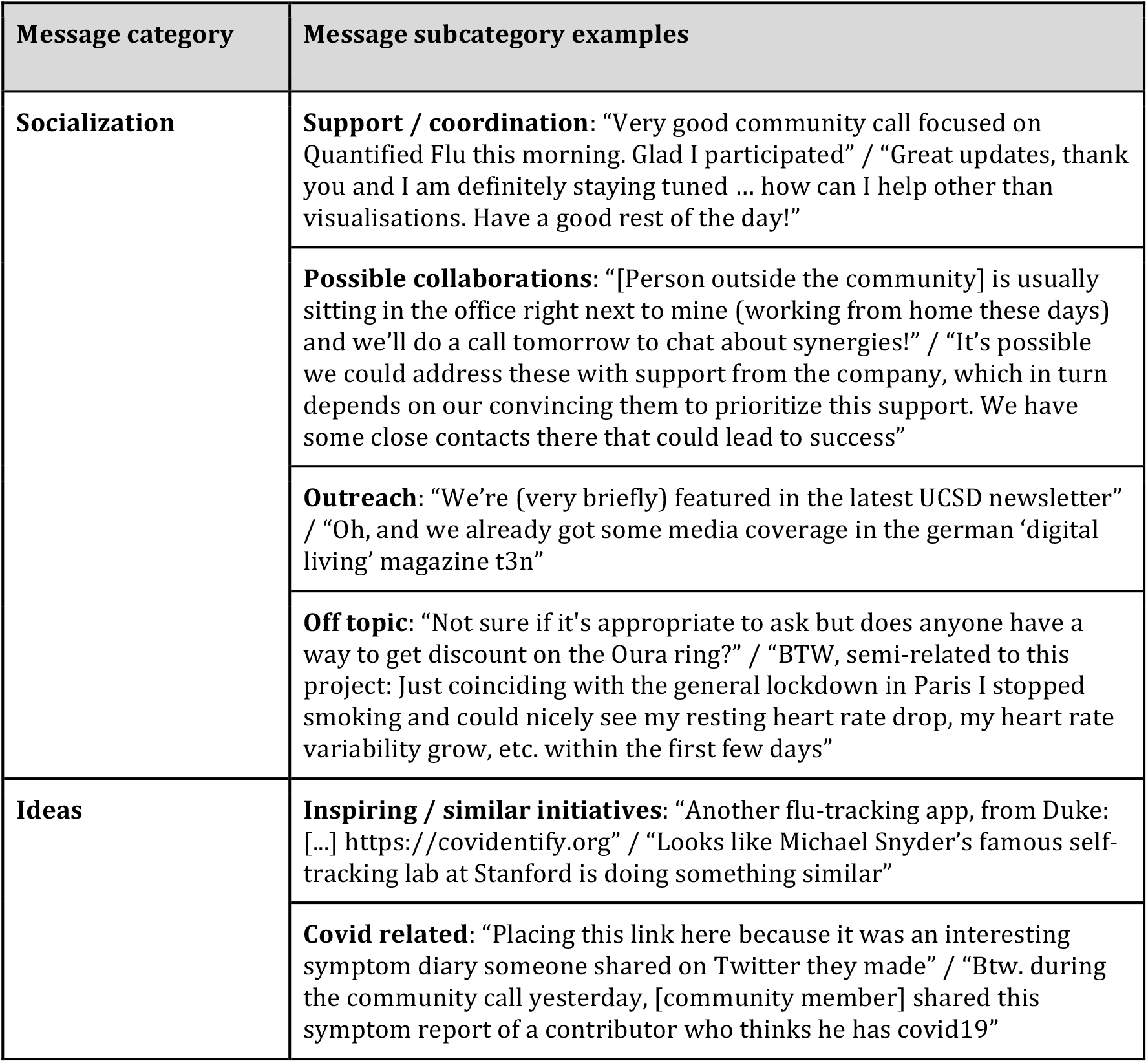
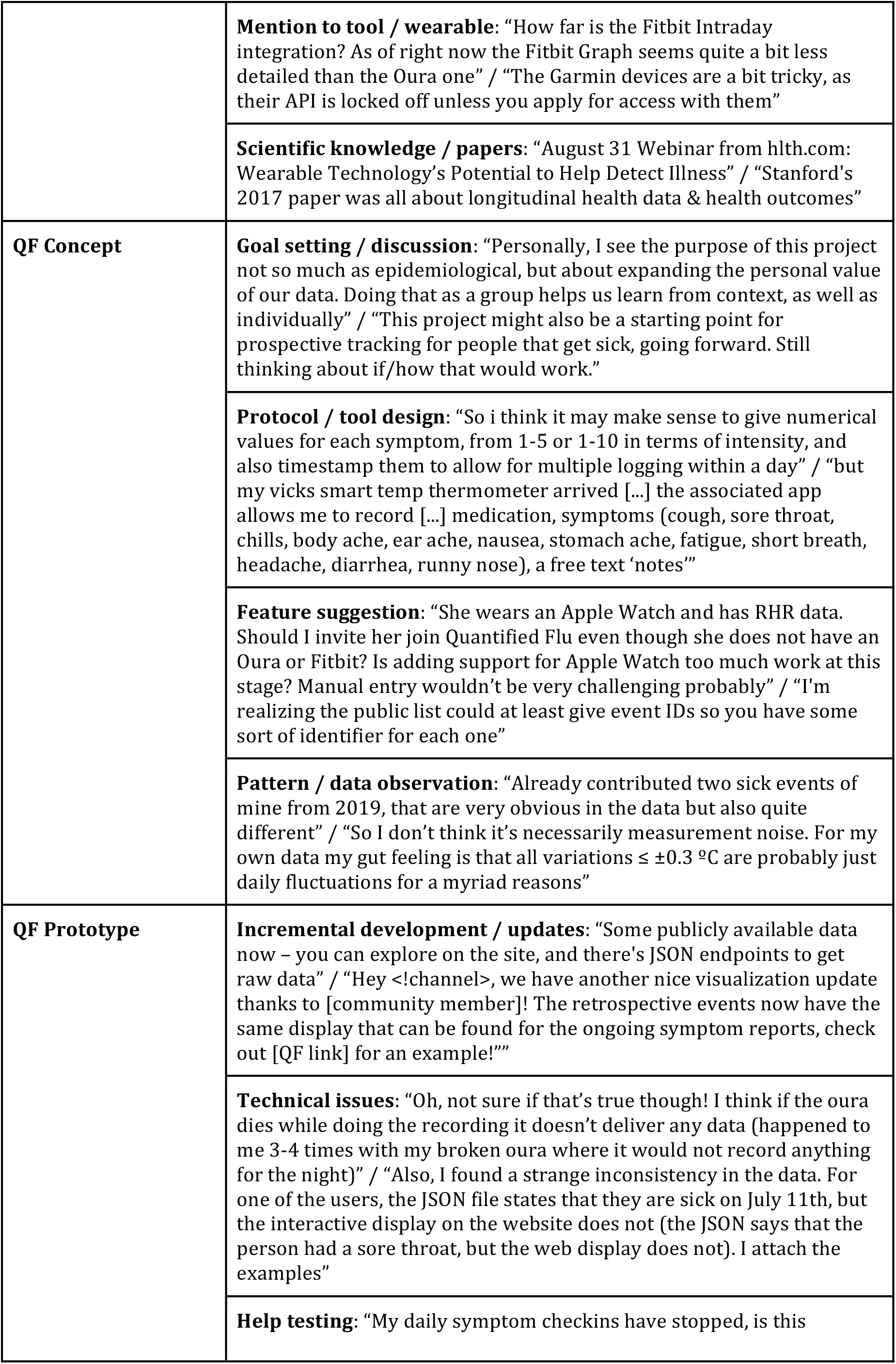
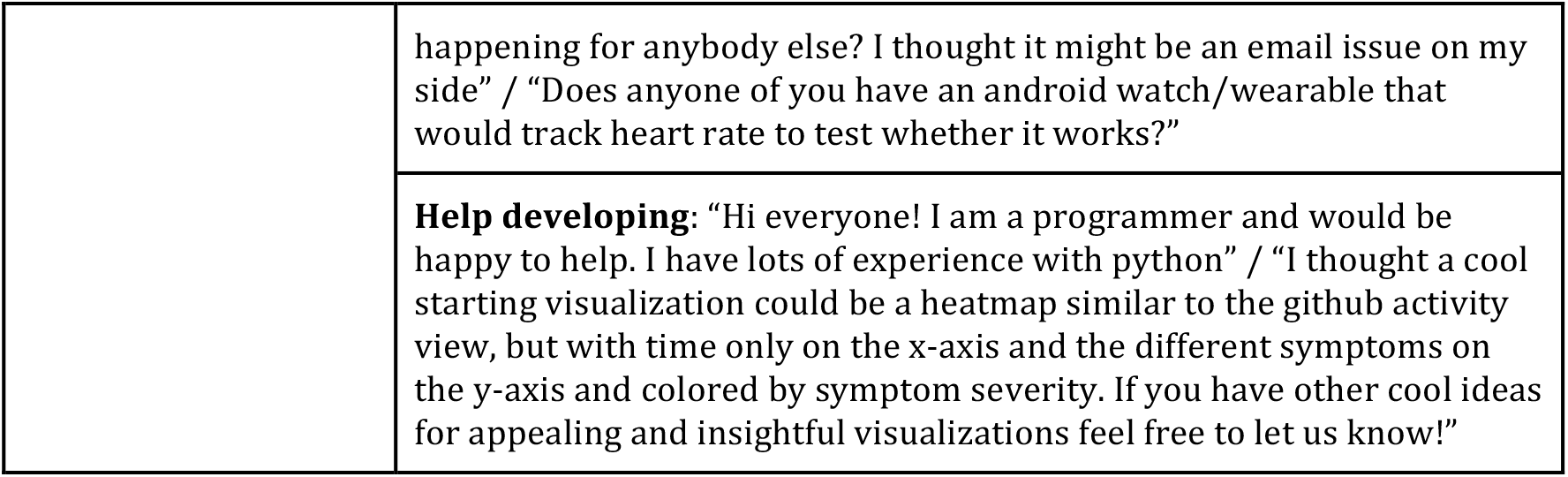
Examples of messages tagged according to the codebook used for the Netnography

Overall, during the development of QF, the *Prototyping* and *Socialization* messages were slightly more common than *Concept* and *Ideas* ones (see bar charts in Figure 1). On the level of the tags or sub-categories, the most frequent ones are *Support / coordination* (227), *Protocol / tool design* (109), *Technical issues* (107) and *Help developing* (106).

**Figure 1:**
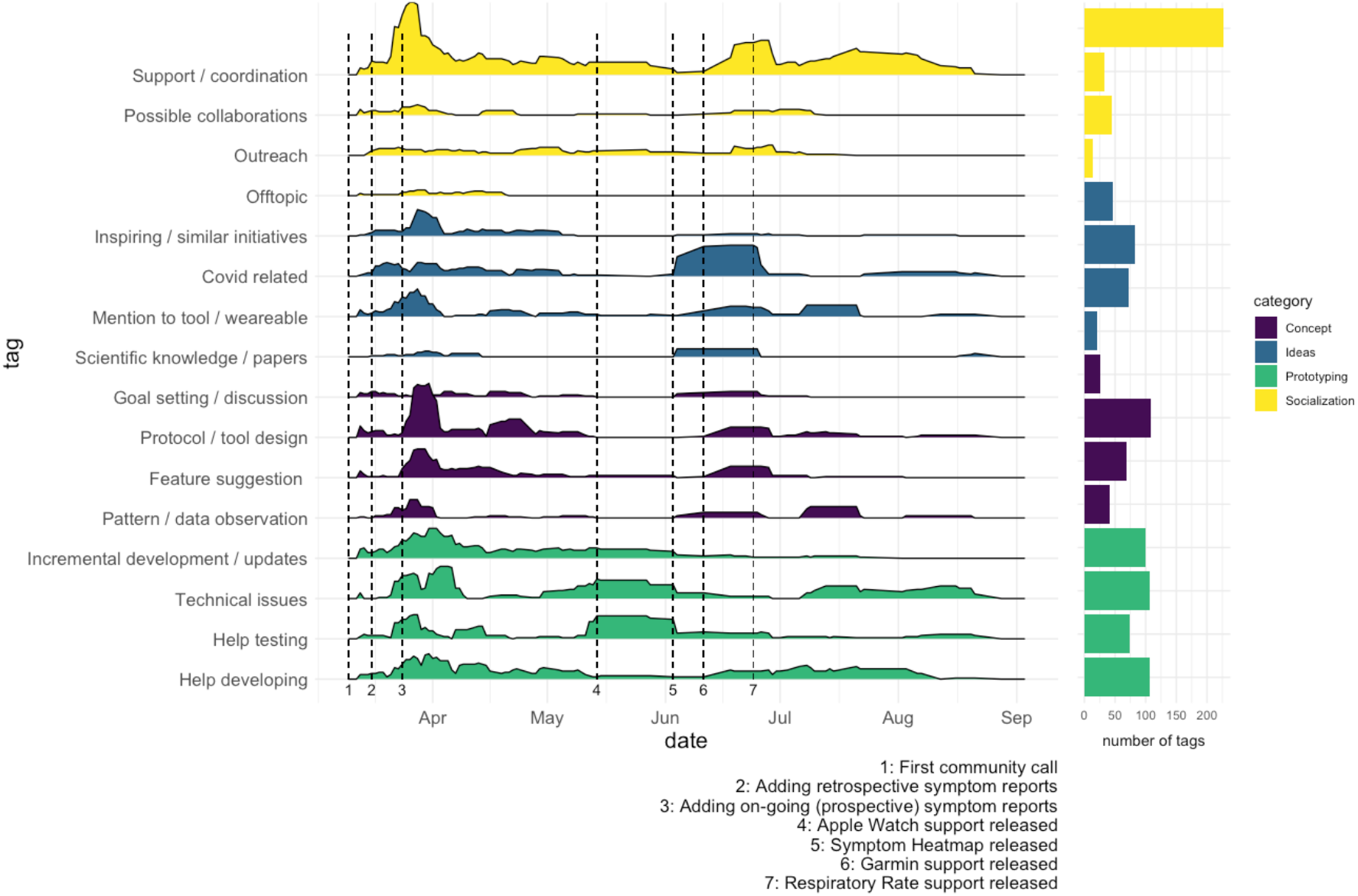
Distribution of message types over time with the frequency of tags are given as 7-day rolling averages. Events 1-7 around QF development are given as vertical lines. Bar plots show the total number of tags per category.

Focusing on these more specific tags, as defined in the codebook (Table 1), within each category over time (Figure 1, left), we observe that all four main categories, as well as the individual tags, are present over the whole time frame of co-creation from early April to September 2020. Particularly, messages regarding *Support / Coordination* are present throughout the whole time range. Other recurrent message types during the analysed timespan fall within the categories *Ideas, Concept* and *Prototyping*, highlighting the iterative design, implementation, and testing participatory processes that took place to develop and improve the QF prototype over time.

Importantly, the *Protocol / tool design, Mention to tool / wearable*, and *Feature suggestions* categories – which are indicative of the co-creation process – do appear early on but remain active in bursts throughout the full observed timespan, often following new releases of the QF prototype. Additionally, the *Help developing* and *Help testing* categories remain active over the whole duration of the prototype development, with the former showing a more constant activity (1.1±1.9 tags per day) while the latter appears in bursts (0.76±2.1 tags per day) around new feature releases.

### Quantified Flu (QF): Technical platform implementation

As a result of this community-based development process, QF evolved into a responsive web application that can connect to a wide variety of devices, implemented in Python/Django programming language. Users must be OH registered users, having an option of linking a range of available wearable devices from which physiological data (heart rate, body temperature, respiratory rate) can be imported into the OH platform; visualize past sickness/infection events (retrospectively) on it (present since the first prototype, launched on March 16, 2020) and engage in daily (prospective) symptom tracking (added in the second prototype, released March 24, 2020) (Figure 2).

**Figure 2:**
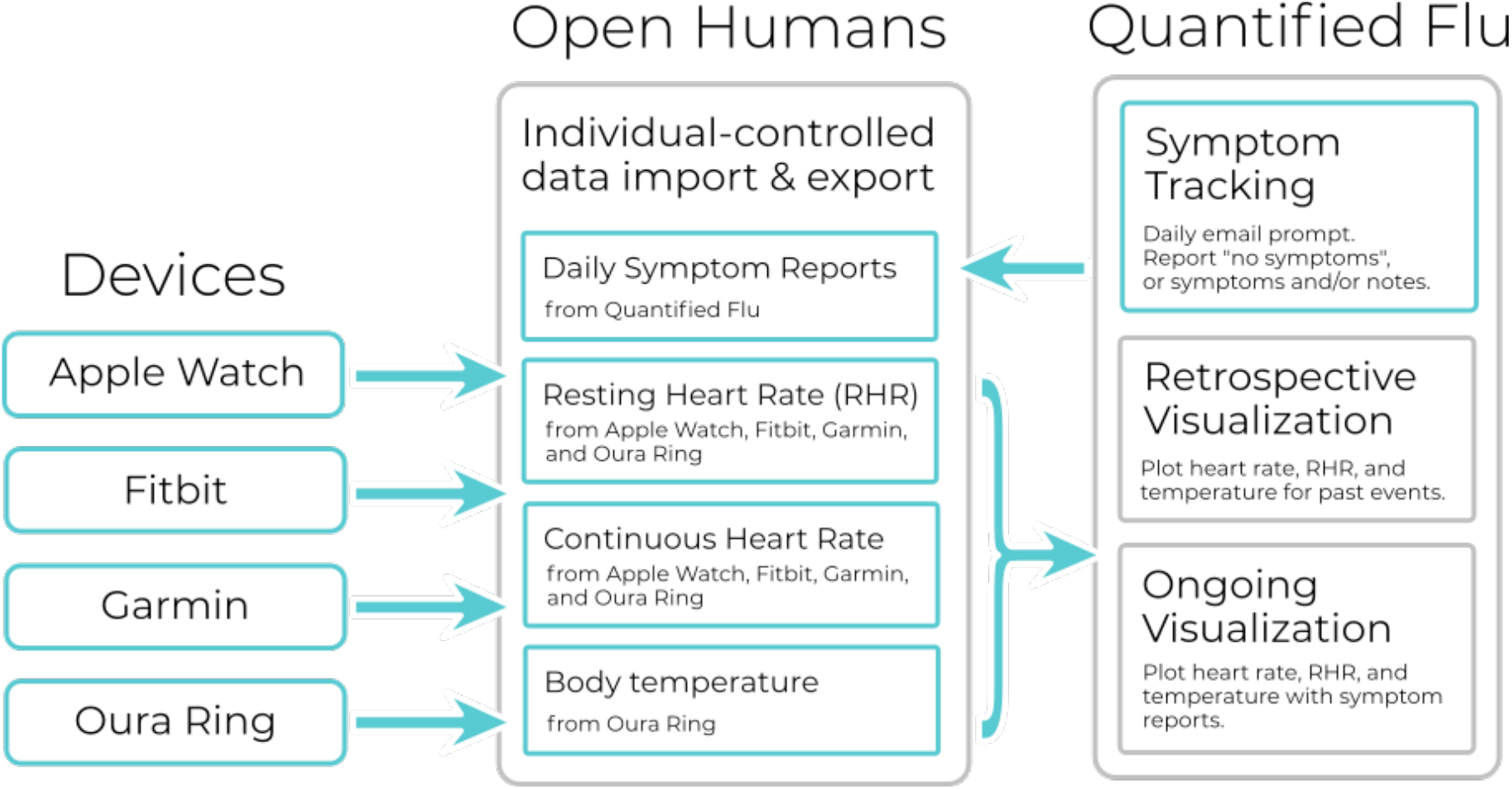
Data- and user-flow in *Quantified Flu*.

#### User accounts, data storing & anonymization

To enable rapid prototyping QF connected to the OH platform [33] as a backend to manage user permissions and store user data. OH provides OAuth2-based APIs to authenticate users while keeping each user pseudonymous to the QF platform, as no personally identifiable information is transmitted. Instead, only a random 8-digit user identifier that is specific to the QF project is provided. Furthermore, OH provides APIs to access and store user data in their system through those identifiers, and gives methods for users to consent to share data from the OH platform with third parties such as QF.

#### Wearables

To further bootstrap the creation of the prototype, QF made use of the existing wearable integrations that OH already offered (Fitbit daily summaries, Fitbit intraday data resolution, and Oura Ring). To facilitate usability QF also integrated those data import methods directly into the prototype, using OH as the data store for the wearable data.

Furthermore, following community suggestions and ideation discussions (see examples in Table 2, “Mention of tool/wearable”), QF also added Google Fit (May 6, 2020), Garmin (June 11, 2020), and Apple Health (May 14, 2020) as additional supported wearable devices. Depending on the wearable, users can import and use their heart rate throughout the day, their resting heart rate, their body temperature, and their respiratory rate in QF (see Table 3).

**Table 3:** Wearables supported by *Quantified Flu*.

| **Wearable** | **Development** | **Resting Heart Rate** | **Heart Rate throughout day** | **Body temperature** | **Respiratory rate** |
| --- | --- | --- | --- | --- | --- |
| **Fitbit** | existing | x |  |  |  |
| **Fitbit intraday** | existing | x | x |  |  |
| **Oura Ring** | existing | x |  | x | x |
| **Google Fit** | extended (added HR data) | x |  |  |  |
| **Apple Health** | added | x | x |  |  |
| **Garmin** | added | x | x |  |  |

Unlike the other wearables integrated into QF, Apple Watch does not provide a web-based API to access and export data. Thus, following another community suggestion (see example in Table 2, “Feature suggestion”) a mobile iOS application was created to provide a link to QF. This specific application enables users to export their heart rate data collected by Apple Watch. The source code for this mobile application is also available under an open license at [43].

#### Symptom tracking

Users can report symptoms through the QF website. Based on prior works [24]–[26] and early community discussion and feedback (see example in Table 2, “Protocol/Tool design”), QF implemented a list of 12 symptoms that are classified as respiratory, gastrointestinal, and systemic symptoms (Table 4), allowing users to score those on a 5-point scale (1=light to 5=worst). Additionally, users can report fever measurements and use free-text fields for the suspected origin of their symptoms, further symptoms, or notes to put their symptoms into context (see example in Table 2, “Protocol / tool design”).

**Table 4:**
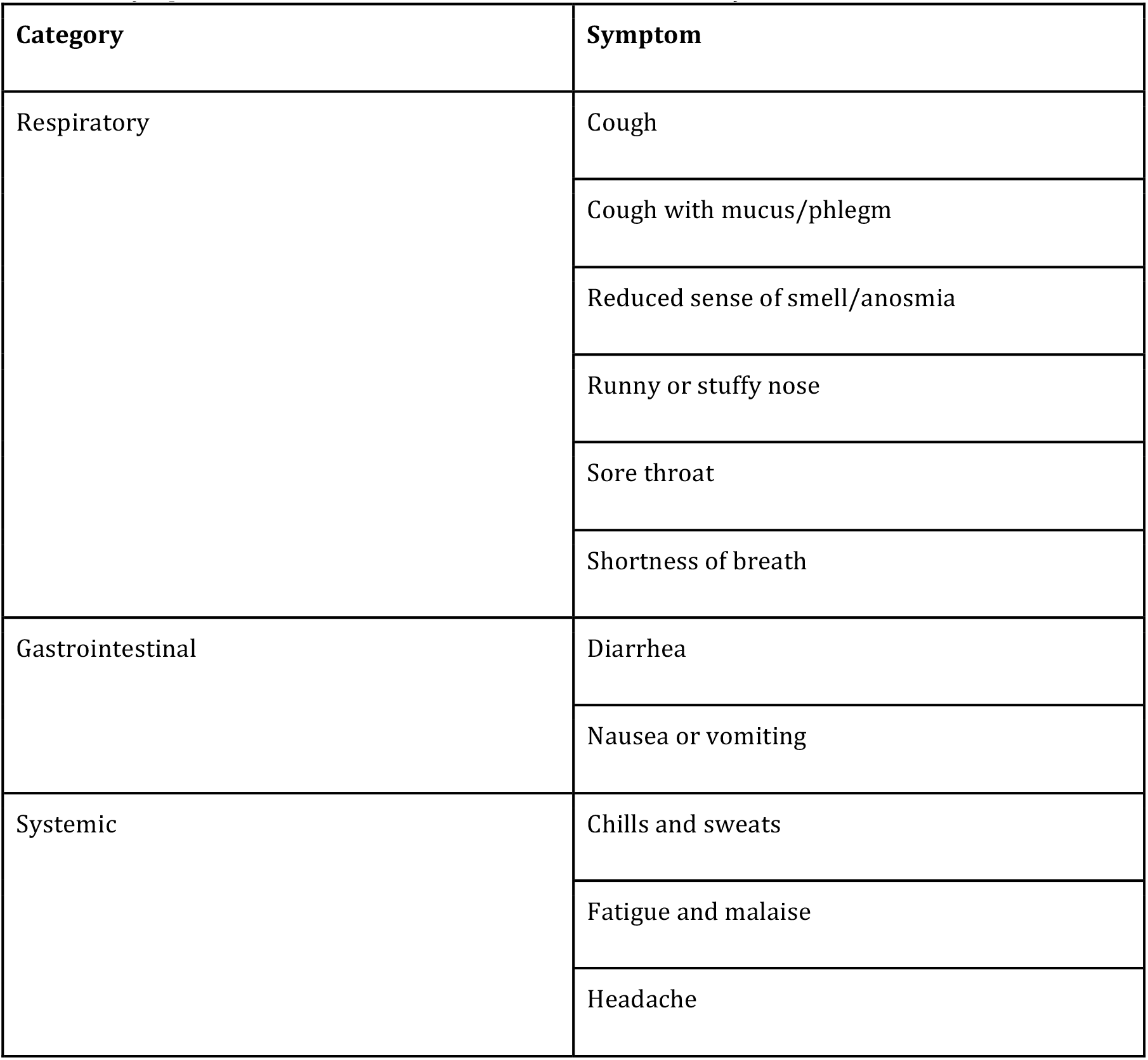
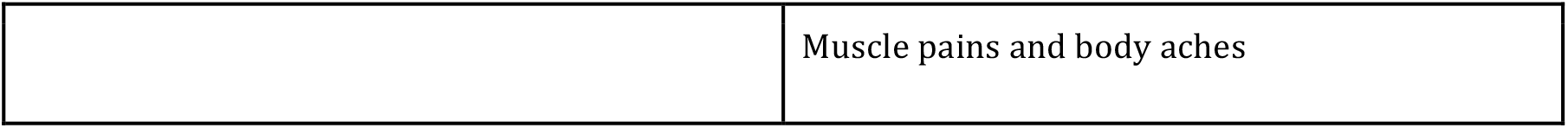
Symptoms of sickness that users can monitor in *Quantified Flu*

Users can opt-in to receive daily symptom report reminders that are sent through the anonymous OH email system at a user-selected time, as another tool feature that was discussed and regularly tested by participants (see example in Table 2, “Help testing”). Each email contains two links: (1) “reporting no symptoms”, single click that requires no further interaction of the user, and (2) “reporting symptoms”, taking users to the symptom report form.

#### Data visualization

To provide users with easy ways to facilitate understanding of their own physiological data, and potentially explore it in relation to their own symptom reports, QF used *D3*.*js* to create interactive visualizations. These visualizations present the evolution of the various physiological data points and put them into the context of their symptom reports where available.

The QF platform gives personal science practitioners two main ways to explore their physiological wearable data in relation to infections: The *retrospective analysis* of prior events as well as an *on-going (prospective) symptom reporting*.

##### Retrospective analysis

Users can select a given, historic date on which they fell sick or have specific symptoms, and QF will – if available – extract wearable data for the 3 weeks prior to that date, as well as 2 weeks after the incident. This allows users to also visualize sickness incidents that happened before the launch of QF. Depending on the wearable (see Table 3), users are given the option to display different physiological variables over that 5 week time period and explore how they change over time. To facilitate the interpretation of changes and outliers in the graphs, both the first and second standard deviations are presented as well (see the screenshot in Figure 3 A). While users can add comments to retrospective events, detailed symptom reports are absent in this mode as most users do not have detailed records of the historic sickness events. The retrospective analyses were part of the first prototype of QF, launched on March 16, 2020.

**Figure 3:**
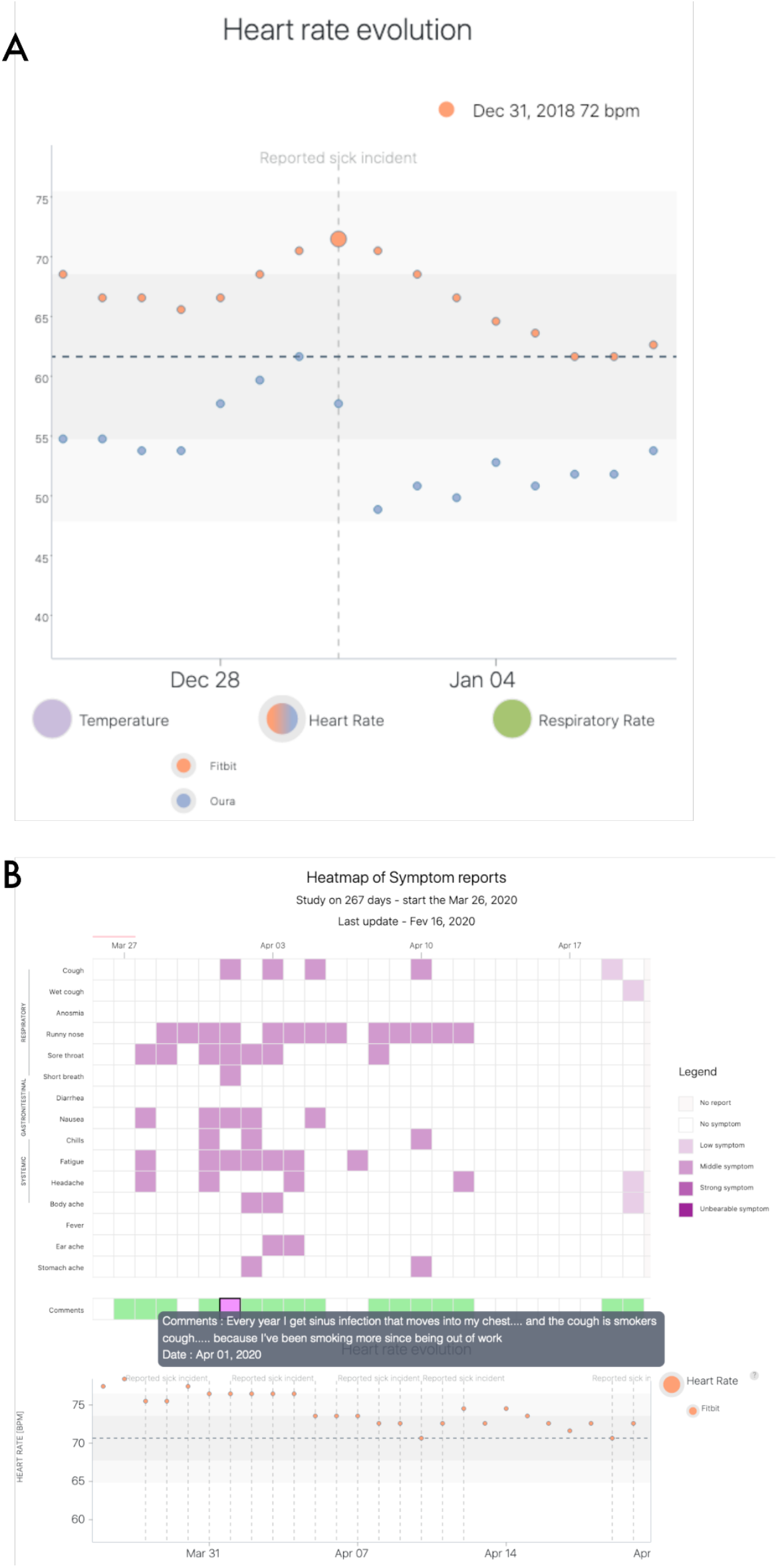
Screenshots of the Quantified Flu prototype website given typical visualizations as generated by users. *(A)* An example data visualization of an individual, retrospective sickness incident that happened on December 31, 2018. Plotted are the resting heart rate recordings as measured by a *Fitbit* and an *Oura Ring. (B)* An example of an on-going symptom report visualization: Top half gives a heat map of which symptoms were present in which strength and green boxes display user-provided free-text comments. The bottom half gives physiological data from wearables.

##### On-going symptom reporting

Users can also report currently experienced symptoms through QF at any moment in time by selecting symptoms and their experienced strengths from a list (see Table 4). This self-report is likely triggered by an email, as explained above. Following symptom reports, users are automatically taken to their data visualization (see screenshot in Figure 3 B). On a wearable device data level, this visualization provides the same details as the retrospective analyses do (see above). The on-going symptom reports (c.f. Figure 3 above) were launched as a new feature in the second iteration of the prototype on March 24, 2020, also following discussion and contributions from the community (see example in Table 2, “Incremental development / updates”).

Additionally, this latter view aligns a heatmap of each daily symptom report to the wearable data timeline, allowing the identification of patterns within the reported symptoms themselves, as well as for visual cross-comparisons between the physiological data and the symptom reports. Furthermore, users can also access their comments for each symptom report from this visualization, allowing them to understand the contexts in which they made those reports.

### Community usage

A total of 190 personal science practitioners have engaged with QF between its launch on March 16, 2020 and December 22, 2020. The initial prototype of QF (in place until March 24, 2020) only offered the possibility to create analyses for retrospective sickness events. This feature was rarely used: Only 24 users tried the feature, creating a total of 47 retrospective analyses. In total, 34 individual wearables have been linked by these 24 users. The prospective on-going symptom report feature was launched on March 24, 2020. In total 92 users made use of this feature at least once, covering a range from a single symptom report being done up to over 300 reports for some members. Overall, 11,658 symptom reports were filed, and 112 wearables have been linked to it, in the time between the feature’s launch and December 22, 2020.

The distribution of user engagement for the whole period (Figure 4, Panel A) – as measured by the number of reports – shows an approximately linear relationship between the number of reports done and the user’s rank of activity. The reports with symptoms are also not equally distributed across all 92 users, with a sizable fraction of users having no or only a few reports that include symptoms, while for some users symptom reports make up half or nearly all of the reports. Overall, the vast majority (10,594 reports, 91 %) were reports that included no symptoms. Of the 1,064 reports with symptoms, 176 (16 %) included explanatory notes or comments in addition to the standardized symptom reports.

**Figure 4:**
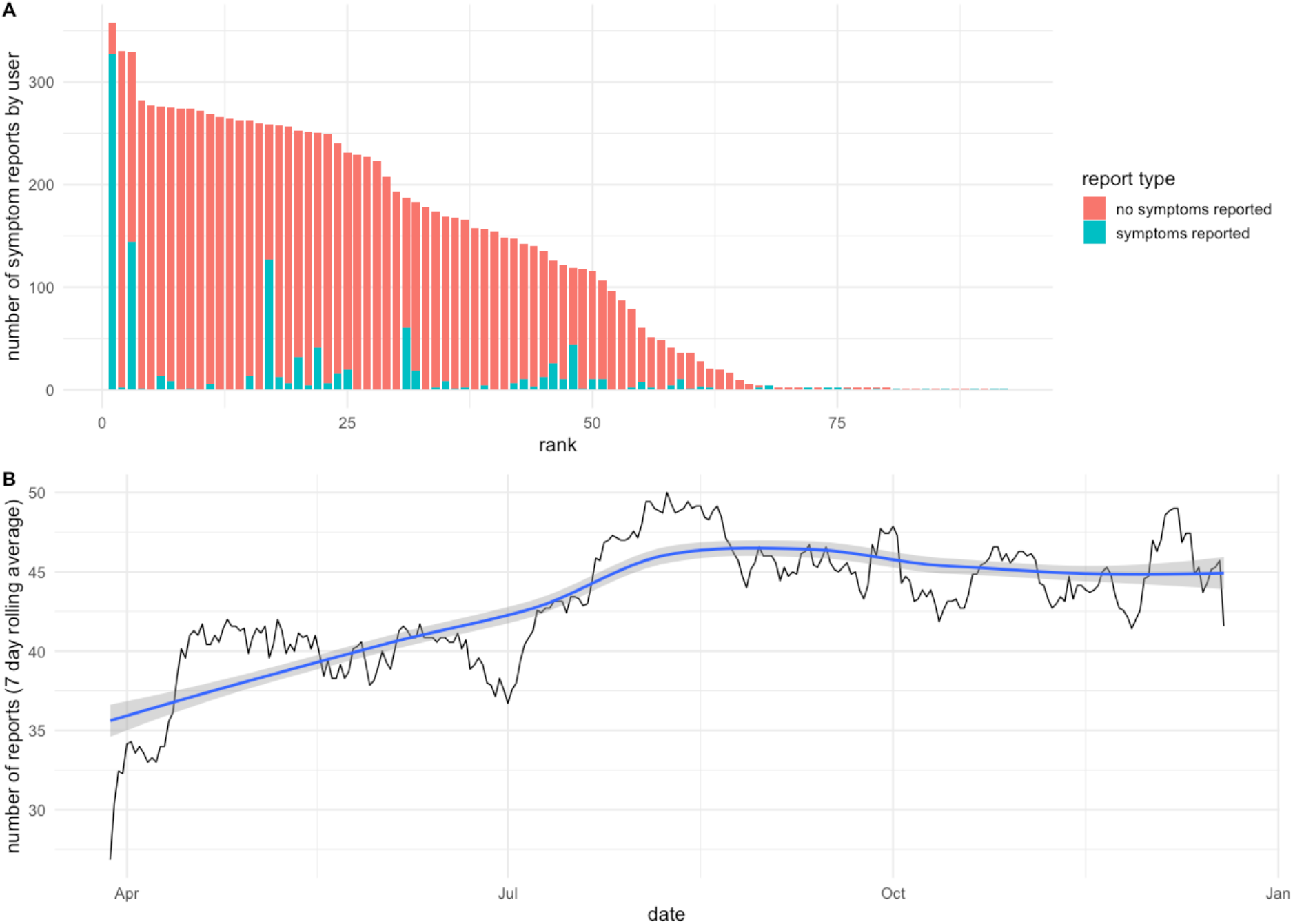
Usage of QF as measured by ongoing symptom reports filed by users. *(A)* Users ranked by the number of symptom reports they have filed, broken down into whether symptoms were reported (blue) or not (red). *(B)* Number of symptom reports filed per day, values were averaged into a weekly rolling average. The blue line gives the loess-smoothed data along with standard error (gray background).

Looking at the number of symptom reports filed per day, we can observe a rapid rise in daily reports at the beginning of April 2020, reflecting the launch of the first prototype with the ongoing symptom reports. The second rise in daily reports happens to start in July 2020, leading to the numbers starting to stabilize at around 45 symptom reports filed per day (by on average 45±5 users per day) (Figure 4, Panel B).

## Discussion

In this paper, we present *Quantified Flu*, a co-created web-based project to enable personal science practitioners to engage with their own wearable data and visualize it in the context of when they are experiencing symptoms of potential infection. The spark that led to this community deciding to co-create a symptom tracking tool was the beginning of the global COVID-19 pandemic, along with population-wide studies that made individuals wonder how useful their own wearables data might be for them in such a pandemic context. With its focus on individual learning, QF stands in contrast to various population-level studies performed to evaluate the usefulness of wearable technology for the prediction of illness [20], [24]–[27]. At this individual scale, symptom tracking and health data more generally can offer support for individual sense-making on health experiences and conditions [44], which can be idiosyncratic and complex [45]. QF was also distinguished by a co-creation approach that targeted the individual learning and research interests of an online community and involved the iterative development of a digital tool in response to feedback, resulting in a format that attracted increased and sustained participation from early users.

One of the main aims of our work was to investigate the consequences of a co-creation approach that focuses on personal science: We observe that the initial QF prototype – which focused solely on retrospective symptom tracking – was rarely used. Only 24 users engaged with this prototype. But importantly, this initial version facilitated additional discussions about designing both the data collection protocol and extending the prototype (see Figure 1), leading to the creation of the on-going symptom reports as a feature launched in the next QF iteration. This feature received much higher attention from the participants, with a total of 92 people using QF for their own regular symptom tracking, delivering some first insight into the importance and potential benefits of early engaging potential users in a health research design co-creation approach.

Furthermore, we observe that the level of engagement across these 92 QF users seems to drop linearly when ordered from most to least engaged users (Figure 4, Panel A). This distribution is untypical for user engagement in online communities, where one typically observes display power-law distributions for engagement [46]. Related to this, digital or mHealth applications in particular typically struggle with achieving continued use, as a large fraction of users drop-out after a few interactions [47], [48]. In previous studies, only 2% of initial users show sustained use in the most extreme cases, with observational studies on average having a 49% dropout rate [49]. In contrast, around half of the QF users that engaged with ongoing symptom tracking did so on a regular basis, leading to 45 symptom reports per day on average (±5), Figure 4 B), and over 50 users reporting more than 3 months of symptom reports, highlighting continued longitudinal use. We argue that these uncharacteristically high numbers of user engagement – which is sustained over time – is a result of the community co-creation process that led to the final prototype of QF. Prior studies have found that users are more likely to continue using mHealth apps if there is a good fit between user and application [50], and in this sense, a co-creation process among future users could be a key way of achieving this fit.

For some users, this continued engagement might furthermore be a sign that they experience regular or recurring symptoms, making them particularly interested in learning empirically about them through this specific kind of self-tracking. This is supported by looking at the number of reports that include symptoms, where a subset of users reports having symptoms frequently, with some users reporting symptoms in 40%, or extreme cases even 90% of the time (Figure 4A). Further evidence for this comes from the notes or annotations that users can submit in the QF website along with their symptoms when filing their daily reports. Looking at the publicly shared notes in these reports, we find examples like “*the cough is smokers cough because I’ve been smoking more since being out of work*” and “*I was deep cleaning the house…all the dust got my allergies going again*”, highlighting possible reasons for recurring symptoms. Furthermore, these annotations help to provide context to individuals and others that aim to re-use publicly shared data: While a severe case of coughing or nasal congestion might hint at acute infection, they might also be unrelated, as the annotations highlight. These contextual descriptions can be hard to formalize, potentially explaining why symptom-based diagnoses are hard to achieve in many cases [51], [52].

Our second main goal, in parallel to the development of the QF prototype itself, was to explore how a community-driven initiative can contribute to collectively creating the tools needed to build self-knowledge by conducting a netnographic analysis of the main QF communication channel. Reflecting on the use of this qualitative and interpretative methodology for the study of online communities [53], we find that it adapts well to user-led prototypes, with some particular strengths and limitations. In the case of QF we find that the netnographic approach was well suited to allow a *post hoc* study of the participatory design process after the prototype creation. This approach could be valuable to get a better understanding of co-creation dynamics in similar health-related projects and studies [54], as it can be applied to existing text corpora of community interactions on digital text tools such as Slack, mailing lists or forums. Its reliance on text communication is also one of the main limitations, as synchronous meetings – virtual and physical – are less accessible as archival data, requiring recordings and transcriptions. Given this, it might be advisable to organize co-creation processes with netnography techniques in mind to ensure adequately sized text corpora.

Applying such a netnographic approach to QF, we found a marked overlap of the various phases of ideation, conceptualization and prototyping over time. While a greater number of interactions can be found in the initial phases, there is a sustained regularity later on, particularly in areas such as feature suggestions or the design of the tool and protocol. In this sense, messages and interactions related to helping with development throughout the whole process reflect a typology of continuous and iterative co-creation, which is typical of collaboration processes in the development of open source tools [55].

This iterative co-creation process is also highlighted in the burst-like appearance of feature suggestions and protocol/tool design discussions, which are frequently appearing following the release of new features, suggesting that new releases spark further protocol refinements and feature ideas, which in turn lead to the QF prototype refinement. Importantly, this means that the protocol itself, along with the concrete implementation, remains in a stage of flux over a longer period of time, compared to more traditional research design approaches. As a result, this type of collaborative approach is at odds with standard ethical oversight procedures for human subject research that require a precise pre-definition of the protocol and the role of the individuals, while a main feature of co-creation is that it is emergent and adaptive, making detailed pre-specifications impossible [56]. To fully take advantage of the benefits of co-creation in participant-led research it might be necessary to develop different models of ethical oversight that recognize the autonomy of participants [57], [58] in order to not discourage or stifle valuable forms of participant-led research [2].

Last but not least, it is also important to highlight how the other types of messages associated with communication in a community of practice context, which favor both online empathy and effective coordination, were produced in a prominent, constant and sustained way from the beginning of the co-creation process (see *Support/Coordination* in Figure 1). This mode of co-creation can be understood as an example of *uninvited citizen science* that relies on a shared set of values, self-stabilizing communication infrastructure, and a loosely defined co-produced knowledge object [59] (e.g., the QF prototype itself). In this way, the development of the data collection platform itself is framed in a dynamic, bottom-up, and adaptive way, similar to other open source and peer production experiences.

## Conclusions

While QF is a project that is still at a prototype stage and with a correspondingly small user base, the co-creation processes of the platform prototype described here represent an example of how the co-development of digital research objects, within the relatively new participatory paradigm of extreme citizen science [60], can be implemented following bottom-up, dialogic approaches and a high level of participant engagement. This aligns with the still scarce literature on what has been called do-it-yourself science or peer-to-peer science [61], [62], in which similar participatory approaches can offer an opportunity for early and sustained engagement from personal science practitioners in the collaborative definition of concepts, features, and protocols for health-related digital platforms.

## Data Availability

All data used in this manuscript comes from public sources and can be accessed under the links provided below.
Additionally all software generated as part of this manuscript is freely accessible under open source licenses in the links provided below.

https://github.com/OpenHumans/quantified-flu

https://quantifiedflu.org/public-data/

http://slackin.openhumans.org/

## Acknowledgements

The authors thank all participants of the Open Humans community calls and the Quantified Self and OpenCovid19 communities for their input and support. We also thank all users of Quantified Flu for their support and feedback. We are also grateful for the suggestions and recommendations of two anonymous reviewers, who helped to substantially improve our manuscript. Thanks to the Bettencourt Schueller Foundation long term partnership, this work was partly supported by the CRI Research Fellowship to BGT. This work was also supported by a microgrant of the OpenCovid19 initiative. KW is supported by the H2020 WellCo project (769765).

## Authors’ Contributions

BGT, KW, GW & MB initiated this study. All authors contributed to the design of the study. BGT, KA, LB, BM and MB performed the software development. BGT, ESH, IB and MF performed data analyses and visualizations. BGT, ESH and MB prepared the original draft of the manuscript. All authors reviewed and edited the manuscript before submission.

## Abbreviations

OH: Open Humans
QF: Quantified Flu

